# Abnormal *TP53* predicts risk of progression in patients with Barrett’s esophagus regardless of a diagnosis of dysplasia

**DOI:** 10.1101/2020.10.18.20213561

**Authors:** Mark Redston, Amy Noffsinger, Anthony Kim, Fahire G Akarca, Diane Stapleton, Laurel Nowden, Richard Lash, Adam J Bass, Matthew D Stachler

**Author notes:** These authors contributed equally to this work (Mark Redston, Amy Noffsinger, Adam J Bass, Matthew D Stachler). **Corresponding Authors** Mark Redston, Dept of Pathology, Brigham and Women’s Hospital, 75 Francis St, Boston, MA 02115, Matthew D Stachler, Dept of Pathology, University of California, San Francisco, 513 Parnassus Ave, San Francisco, CA 94143. **Author Contributions:** Data analysis: MR, MDS, AN; Computational analysis: MDS; Patient selection and clinical data collection: MR, AN, DS, LN; DNA isolation: FGA, AK; Histologic review: MR, AN, RL, MDS; Conceived of study: MR, AN, RL, MDS, AJB; Manuscript preparation: MDS, MR; Manuscript review: MDS, MR, AN, AK, FGA, RL, AJB. Disclosures: The authors declare no competing financial, personal, other interests.

## Abstract

Barrett’s esophagus is the precursor to esophageal adenocarcinomas, which are deadly cancers with a rapidly rising incidence. A major challenge is identifying the small group with Barrett’s esophagus who will progress to advanced disease from the many who will not. Assessment of p53 status has promise as a predictive biomarker, but analytic limitations and lack of validation in large, definitive studies have precluded its use. In this study, criteria for abnormal immunohistochemical expression of p53 were developed in non-dysplastic Barrett’s biopsies and validated with sequencing to assess *TP53* mutations. The utility of p53 expression as a biomarker for progression of Barrett’s esophagus was tested retrospectively in 561 Barrett’s patients with or without known progression. The findings were prospectively validated in a clinical practice setting in 1487 Barrett’s patients. Abnormal p53 expression highly correlated with *TP53* mutation status (90.6% agreement) and strongly associated with neoplastic progression in the retrospective cohorts, regardless of histologic diagnosis (P<0.001). In patients with non-dysplastic Barrett’s esophagus at baseline, 89/179 (49.7%) of those with subsequent progression to high grade dysplasia/cancer and 3/179 (1.7%) of non-progressors had abnormal p53 (sensitivity: 49.7,% specificity: 98.3%, Odds ratio: 58 (95% CI 17.9-188.5, P<0.0001) for identifying progressors). In the prospective validation cohort, p53 immunohistochemistry predicted progression among non-dysplastic Barrett’s, indefinite for dysplasia, and low-grade dysplasia (P<0.001). p53 immunohistochemistry can successfully identify Barrett’s esophagus patients at high risk of progression, including in patients without evidence of dysplasia. p53 immunohistochemistry is inexpensive, easily integrated into routine practice, and should be considered in biopsies from all Barrett’s patients without high grade dysplasia or cancer.

## INTRODUCTION

Esophageal adenocarcinoma (EAC) is an increasing cause of both morbidity/mortality and healthcare burden given its dismal five-year survival rate and its striking increase in incidence.^1,2^ EAC arises from a pre-neoplastic precursor, Barrett’s esophagus (BE), which forms in response to reflux injury to the lower esophagus. The prevalence of BE in the US population has been estimated to be from 1.6% to 10% of adults, putting millions at heightened risk of EAC.^3,4^ Patients with BE are currently recommended to undergo frequent endoscopic surveillance wherein biopsies are evaluated for histopathologic signs of progression, namely dysplasia. However, the annual incidence of progression among patients with BE remains low, estimated at ≤0.33%.^5^ Coupled to the large population with BE, this low risk of progression challenges the feasibility and cost-effectiveness of screening. An improved method of identifying which patients are at increased risk of progression, especially in the large population of those with nondysplastic BE, would greatly facilitate developing more effective surveillance and treatment strategy.

Currently, the presence of dysplasia based upon pathologic assessment of BE biopsies is used to select patients for more intense screening, ablation/endoscopic mucosal resection, or rarely even surgical esophagectomy. However, there are limitations to relying on dysplasia as the sole sign of increased cancer risk. First, there is substantial inter-observer variability in histological grading of dysplasia.^6–9^ Second, we lack accepted biomarkers to identify at-risk patients among the large population of patients with non-dysplastic BE (NDBE). Furthermore, evidence now suggests that the progression from dysplasia to cancer can occur more rapidly than originally thought^10^ indicating that current surveillance strategies focused upon finding patients in the window of time between onset of dysplasia and development of advanced cancer may be ineffective. Improved methods to identify high risk individuals with BE prior to the onset of dysplasia may thus both enhance the efficacy of screening and provide economic value by focusing resources on the minority of BE patients who may ultimately progress.

In attempts to improve patient stratification, previous biomarker studies have queried mutations, chromosomal alterations, copy number/aneuploidy^11–13^ and methylation of specific genes.^14^ While advanced molecular diagnostics are promising routes to biomarker development, many studies have utilized testing strategies that are difficult to translate into routine clinical practice. Until robust and cost-effective molecular diagnostics can be developed, there is an immediate need for biomarkers that are inexpensive, can be quickly adopted by current clinical laboratories, and can be readily interpreted by pathologists. Mutation or aberrant expression of tumor suppressor p53 has been identified as a candidate risk factor for progression in BE, but clinical use of p53 immunohistochemistry (IHC) has been largely limited to supporting the diagnosis of dysplasia.^15–18^ Additionally, many studies evaluating p53 IHC focused on overexpression only, disregarding the absent staining pattern which has been shown to be an important component as it may mark tissues with truncating mutations of the gene^18,19^. Recent molecular studies specifically focusing on NDBE tissue taken years prior to progression to advanced disease identified *TP53* mutations in over 45% of such samples and was associated with a 16-fold increase risk of progression.^20–23^ While these new genomic data suggest the potential value of p53 testing in routine surveillance, p53 IHC is currently not recommended for risk stratification.^24,25^ We sought to determine the applicability of p53 IHC to risk-stratify patients with NDBE, Barrett’s esophagus indefinite for dysplasia (BE-IND), or Barrett’s esophagus with low-grade dysplasia (BE-LGD) and identify those most likely to progress to either BE-HGD or EAC using a large, representative collection of routine screening and surveillance biopsies taken at community endoscopy centers throughout the United States.

## RESULTS

### Patient selection and cohort design

After IRB approval, the pathology records of Inform Diagnostics (Irving, Texas), a national GI pathology laboratory providing services to gastroenterologists throughout the US, were searched from 2001 to 2019 to identify patients who were undergoing endoscopic surveillance for Barrett’s esophagus. Three separate groups of patients were identified for study (**Figure 1**). First, as detailed in the **Methods**, an initial cohort was selected to determine and validate p53 IHC grading criteria.

**Figure 1:**
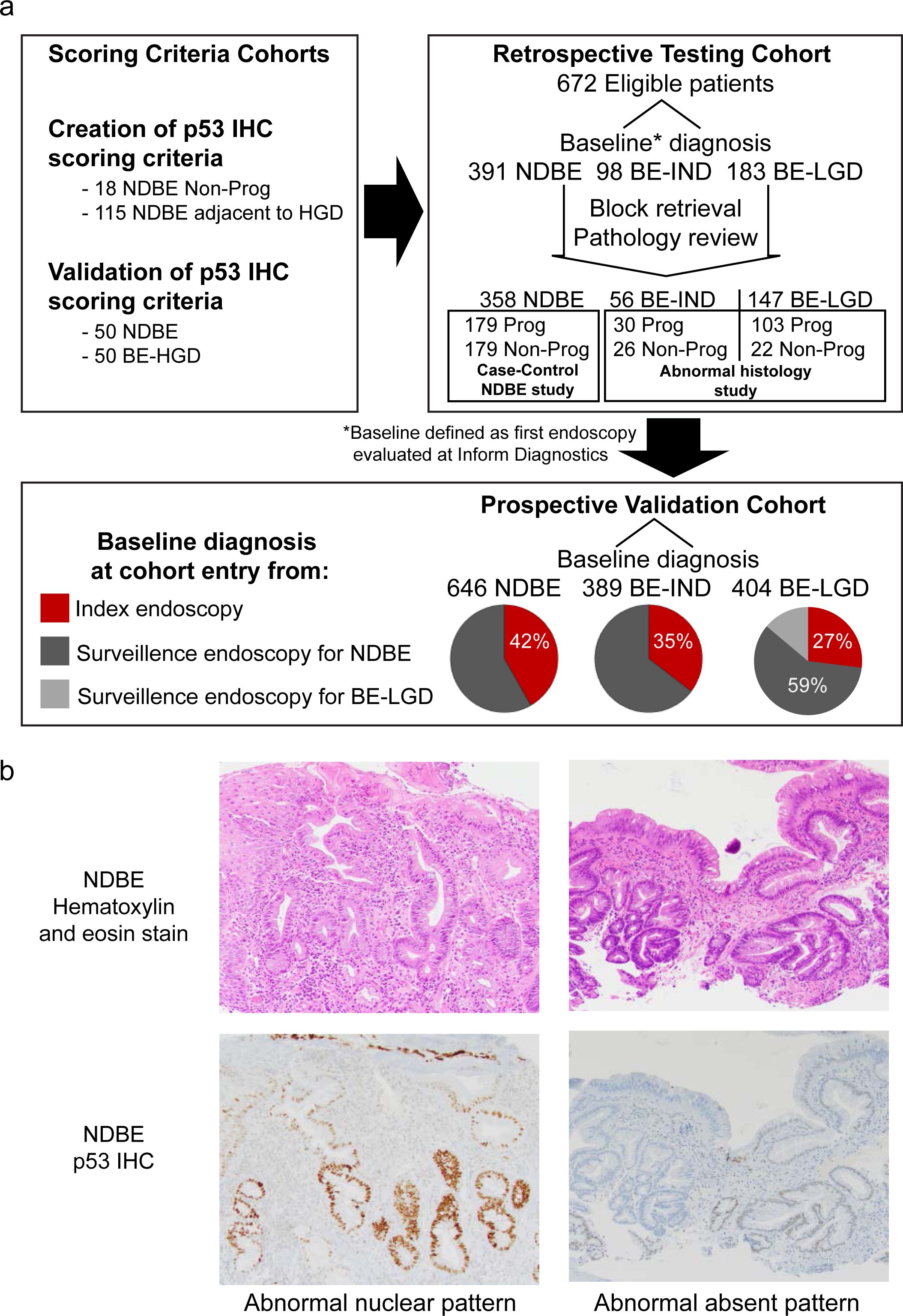
Barrett’s Samples and p53 IHC staining. a) Diagram of the samples used for the creation of IHC scoring criteria, testing cohorts, and validation cohorts. b) Example photomicrographs of hematoxylin and eosin or p53 immunohistochemistry stained slides.

Second, we searched retrospectively for patients with known follow-up (at least two separate endoscopic surveillance biopsies) between 2001 and 2011. These patients were divided into progressors (all available patients having a baseline diagnosis of NDBE, BE-IND, or BE-LGD followed by a diagnosis of BE-HGD or EAC) and non-progressors (patients having a baseline diagnosis of NDBE, BE-IND or BE-LGD with at least 3 years follow up without progression confirmed by at least 1 additional endoscopy with biopsies, **Methods)**. Baseline endoscopies were defined as the first endoscopy with a diagnosis of Barrett’s esophagus seen at Inform diagnostics. In order to determine the ability of p53 IHC to identify patients with NDBE at risk of progression, a case-control cohort of patients with an initial baseline diagnosis of NDBE and endoscopic follow-up was constructed (**Figure 1, Table S2**). After exclusions (See **Tables S2-S4**) there were 179 patients with available NDBE samples taken prior to subsequent progression to HGD or EAC and 179 non-progression patients with NDBE who were matched on patient age and gender. Additionally, 56 patients with a baseline diagnosis of BE-IND (30 progressors and 26 non-progressors) and 147 patients with a baseline diagnosis of BE-LGD (103 progressors and 44 non-progressors) were identified and studied as a separate cohort of patients with abnormal histology. This cohort included all identified progressing and non-progressing BE-IND and BE-LGD patients with appropriate follow up. All samples underwent central pathologic review of sections cut concurrently with the IHC slides to confirm the diagnosis. All morphologic review was blinded to outcome and p53 IHC status, and all p53 IHC review was blinded to outcome and morphologic diagnosis. Clinical characteristics of subjects are summarized in **Table 1**.

**Table 1.** Clinical characteristics of subjects in retrospective testing cohorts.

|  | NDBE at baseline |  | BE-IND at baseline |  | BE-LGD at baseline |  |
| --- | --- | --- | --- | --- | --- | --- |
|  | Non-progressor | Progressor | Non-progressor | Progressor | Non-progressor | Progressor |
| *Total* | 179 | 179 (38 EAC) | 26 | 30 (3 EAC) | 44 | 103 (16 EAC) |
| *Age (years)* |  |  |  |  |  |  |
| Median | 67 | 66 | 65 | 62 | 68 | 66 |
| Mean | 66.0 | 65.7 | 67.3 | 63.6 | 68.4 | 65.7 |
| Range | 38-92 | 41-87 | 47-80 | 45-84 | 43-82 | 40-85 |
| *Gender* |  |  |  |  |  |  |
| Female (%) | 25 (14.0%) | 25 (14.0%)<sup>a</sup> | 6 (23.1%) | 1 (3.3%)<sup>b</sup> | 14 (31.8%) | 16 (15.5%)<sup>b</sup> |
| Male (%) | 154 (86.0%) | 154 (86.0%) | 20 (76.9%) | 29 (96.7%) | 30 (68.2%) | 87 (84.5%) |
| *Follow-up (years)<sup>c</sup>* |  |  |  |  |  |  |
| Median | 5.37 | 2.26 | 4.55 | 1.04 | 4.81 | 0.9 |
| Mean | 5.78 | 2.98 | 5.30 | 1.54 | 5.28 | 1.47 |
| Range | 3.07 - 13.68 | 0.09-9.9 | 3.29 - 9.02 | 0.21 - 4.72 | 3.03 - 11.14 | 0.06 - 11.06 |
| *Total accessions per subject* |  |  |  |  |  |  |
| Mean | 3.13 | 2.73 | 3.58 | 2.6 | 4.48 | 2.45 |
| Median | 3 | 2 | 3 | 2 | 4 | 2 |
| Range | 2 - 9 | 2 - 9 | 2 - 6 | 2 - 4 | 2 - 11 | 2 - 8 |
| *Baseline Pathology blocks per subject* |  |  |  |  |  |  |
| Mean | 1.23 | 1.51 | 2.04 | 1.57 | 2.02 | 1.91 |
| Median | 1 | 1 | 1 | 1 | 1 | 1 |
| Range | 1 - 6 | 1 - 7 | 1 - 8 | 1 - 5 | 1 - 10 | 1 - 8 |

<sup>a</sup> Negative at baseline subjects were gender matched.
<sup>b</sup> Fisher exact P<0.05.
<sup>c</sup> Selection criteria for subjects were such that follow-up was longer in non-progressors than progressors.

The third group of patients was used to validate the findings from the retrospective studies and determine how p53 IHC may be utilized to prospectively stratify outcome in a clinical setting. As part of a quality improvement project and after collective review sessions, the p53 IHC scoring criteria developed in this study were adopted by the Inform Diagnostics GI pathology team for standardized reporting of p53 IHC on the pathology reports and was implemented in June 2011 in routine diagnostics. Pathology records from 2011 to 2019 were searched to identify patients using the following criteria: i) a biopsy diagnosed as NDBE, BE-IND, or BE-LGD; ii) a p53 IHC result reported in the original pathologic diagnosis; and iii) at least one follow-up esophageal biopsy. To determine how p53 IHC could be implemented in practice, analyses were performed using data from the original pathologic diagnosis without central pathology review. However, all diagnoses of progression were reviewed to confirm the diagnosis. Immunohistochemical staining was performed at 3 different laboratories and over 40 different pathologists interpreted the p53 IHC.

### Development of p53 immunohistochemistry scoring criteria and correlation with *TP53* mutations status

We first developed our p53 IHC scoring by performing IHC staining on 18 NDBE biopsies from patients with no known dysplasia and in 115 NDBE biopsies from patients with concurrent HGD as the latter cases were most likely to have p53 abnormalities. Scoring criteria (**Methods, Table S1**) were selected to show 100% specificity (IE 0/18 NDBE from patients without dysplasia were positive), which yielded 39/115 (34%) positive NDBE biopsies in patients with concurrent HGD. The staining criteria was validated using 50 unselected NDBE biopsies and 50 BE-HGD biopsies. Abnormal p53 staining was seen in 2/50 (4%) of the NDBE biopsies and 48/50 (96%) BE-HGD biopsies, confirming the scoring criteria are both sufficiently sensitive and specific.

To evaluate how p53 IHC status relates to *TP53* mutation status we next performed *TP53* sequencing on a subset of samples. We were able to obtain DNA for adequate sequencing from 92 BE samples derived from 28 progression patients and 6 non-progression patients. *TP53* mutations were identified in the DNA from 50 of these samples, specifically from 21 progression patients and 3 non-progression patients. In 83/92 samples (90.2%) the mutational status correlated with the p53 IHC results. There were 2 samples in which a *TP53* mutation was called but IHC was read as normal and there were 7 samples that were negative for a mutation but read as abnormal with IHC. Interestingly, of the 7 samples negative for mutation and abnormal by IHC, 6 had an absent IHC staining pattern, including 5 samples from a single patient, raising the hypothesis of an alternative pathway of p53 silencing in that patient. In samples with both abnormal p53 IHC and a *TP53* mutation, all 10 samples with an absent p53 IHC pattern had a mutation that would likely lead to a truncated protein, confirming the importance of recognizing the absent pattern as a marker for these pathogenic *TP53* mutations. All but 1 sample with strong nuclear p53 IHC staining had a missense mutation recurrently seen in cancer. In total, p53 IHC was 96% sensitive and 83.3% specific for identifying *TP53* mutations in the sequenced samples.

### Baseline p53 immunohistochemistry status predicts progression regardless of histologic diagnosis

We performed p53 IHC on the case-control cohort of NDBE and the patients with BE-IND or BE-LGD (**Figure 2**). p53 IHC grading was performed blinded to progression status. Biopsies from progressors were much more likely to be p53-ABNL than biopsies from non-progressors. In patients who progressed to advanced disease, p53 positivity in baseline endoscopies (**Figure S1**) was 89/179 (49.7%), 27/30 (90.0%), and 97/103 (94.2%) in NDBE, BE-IND, and BE-LGD respectively. These numbers were dramatically lower in non-progressing patients with 3/179 (1.7%), 4/26 (15.4%), and 20/44 (45.4%) positive in NDBE, BE-IND, and BE-LGD respectively (P<0.00001 for all) (**Table S6 and S7**). **Figure 2** shows accession timeline charts of diagnoses and p53 IHC in each biopsy over time for all non-progressors and progressors, grouped by baseline histology, and sorted by duration of follow-up. The results were similar when analyses were preformed using the original pathologic diagnosis (rather than the central pathology review) (**Tables S8 and S9**).

**Figure 2:**
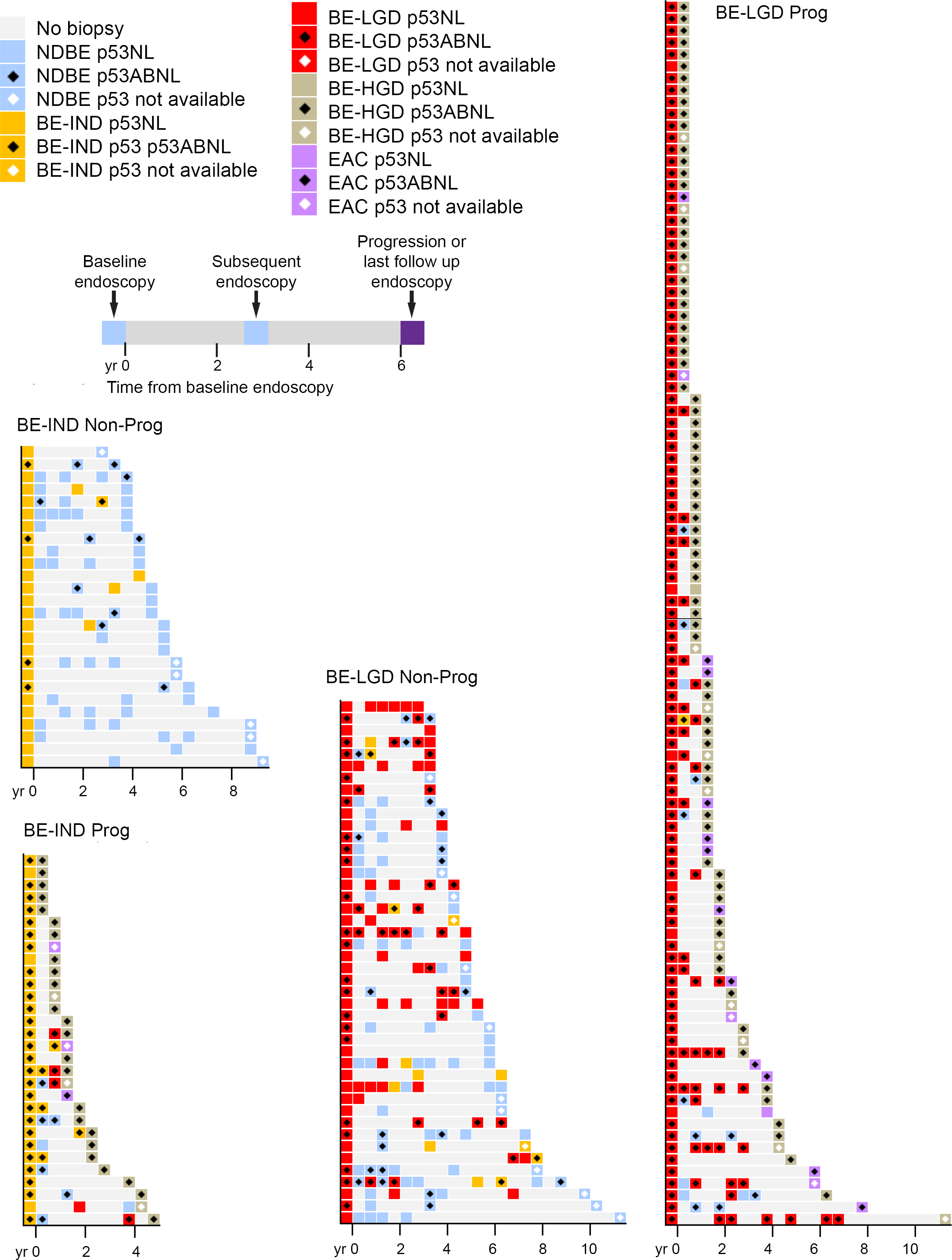
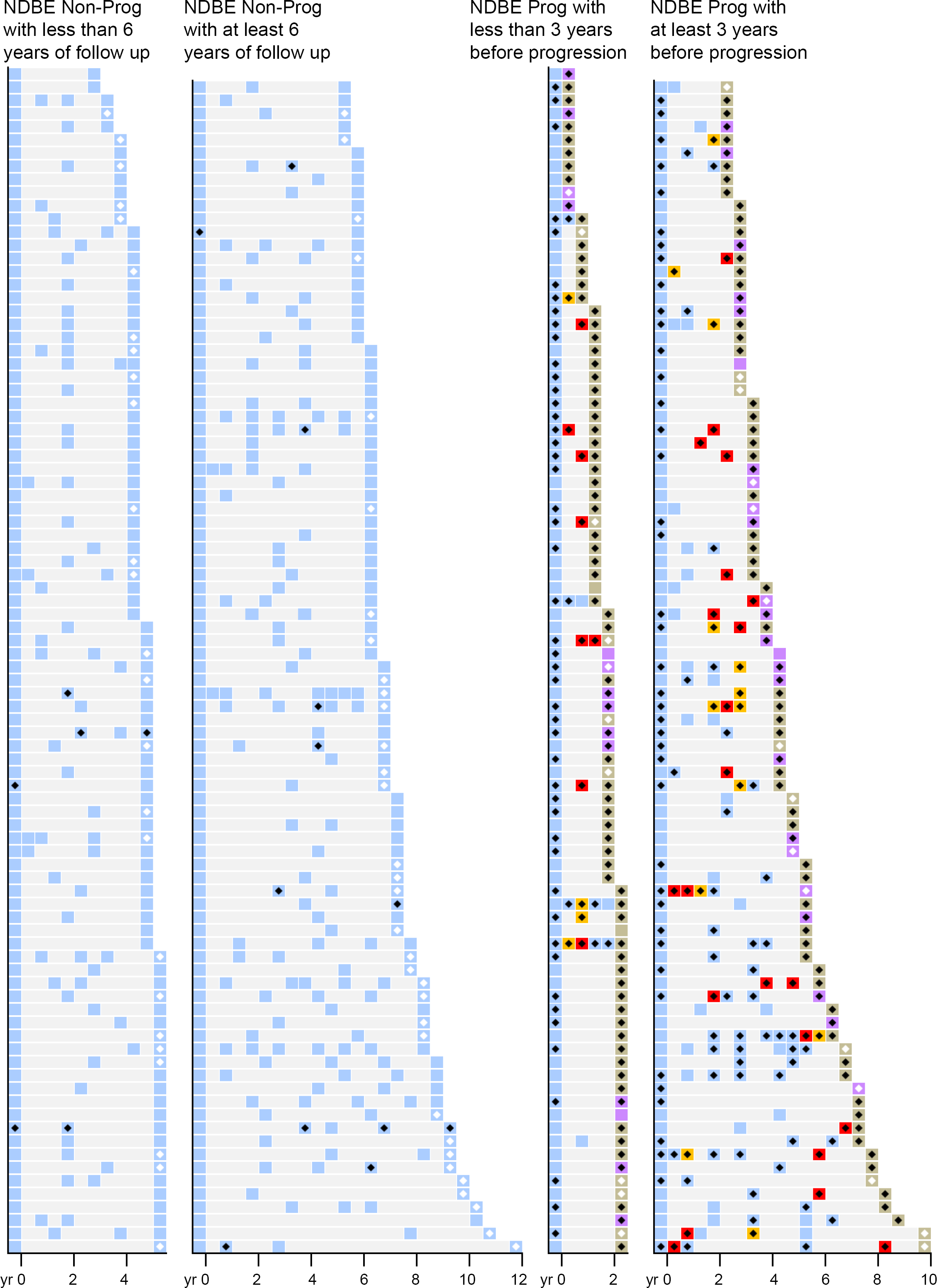
p53 IHC positivity in retrospective testing cohort. Charts showing timing, histologic diagnosis, and p53 IHC positivity for all patients with a baseline of NDBE, BE-IND, or BE-LGD diagnosis separated by progression status. Each row represents a single patient and timeline on x-axis shows when endoscopies occurred with baseline endoscopy at time 0.

### p53 positive non-dysplastic Barrett’s esophagus biopsies are highly enriched in patients who will progress

In our NDBE case-control cohort, there were 24 progression subjects who had a p53-NL baseline endoscopy and had later surveillance endoscopies with NDBE before any higher-grade diagnosis (initial light blue box with additional box/sample before progression, **Figure 2B**). In these 24 subjects, the follow-up was p53-ABNL in 19/38 (50.0%) of the NDBE biopsies from 14/24 (58.3%) subjects (**Figure S2**). Combined with the original baseline NDBE results (**Tables S6 and S7**), a total of 131/317 (41.3%) NDBE biopsies and 103/179 (57.5%) subjects were p53-ABNL prior to the development of progression, IND, or LGD. In contrast, if all baseline and follow-up/surveillance biopsies are combined in NDBE non-progressors, p53-ABNL was only present in 18/620 (2.9%) biopsies and 14/179 (7.8%) subjects (Chi square P<0.00001 for both). Importantly, 8 of these latter 14 subjects were only p53-ABNL in biopsies in the latter part of their follow-up, with 3 years or less subsequent follow-up since becoming p53 positive (**Figure 2B**). Therefore, it is unknown if these patients will eventually progress.

We then limited our NDBE cohort to only subjects who exclusively had a NDBE diagnosis throughout surveillance and who had biopsies greater than one year before progression to determine the rate of p53 positivity in this most restricted population. In this group of clinically and histologically very low risk subjects, p53 IHC in baseline endoscopies identified 60/127 (47.2%) of progression subjects compared to only 3/179 (1.7%) of non-progressors (P<0.0001). When all NDBE biopsies/endoscopies were considered, the rate of p53 positivity increased to 72/127 (56.7%) of progressors and 14/179 (7.8%) of non-progressors (P<0.0001).

### p53 IHC allows identification of higher risk patients earlier and more frequently than a diagnosis of low-grade dysplasia or indefinite for dysplasia

As the current standard is to use histologic abnormalities (i.e. dysplasia) to define high-risk features, we evaluated the pre-progression biopsies in all patients with subsequent progression to determine when p53 positivity emerged relative to the detection of a histologic abnormality. The prevalence of p53-ABNL was calculated for sequential time points prior to progression and was found to be table over time (**Figure 3, Supplementary Figure S3**). The only notable drop of p53-ABNL occurred in the small subgroup of NDBE biopsies taken more than 7 years prior to progression [5/18 (27.8%) p53-ABNL, Chi Squared NS compared to NDBE biopsies within 7 years of progression 161/315 (51.0%)]. In contrast, a diagnosis of BE-IND or BE-LGD occurred closer to progression with the prevalence of histologic abnormalities (BE-IND or BE-LGD) in all progressor biopsies falling steadily between 1-3 years prior to progression (**Figure 3**). Abnormalities in p53 IHC were present at a higher frequency than abnormalities in morphologic diagnosis at all timepoints, and this difference was most striking at time points more than 2 years prior to progression, again highlighting the potential value of p53 testing in the NDBE population. Importantly, at 3-5 years prior to progression, which encompasses the current surveillance guideline of Barrett’s patients with no prior dysplasia, morphologic abnormalities were found in only 23/87 (26.4%) of endoscopies in patients with subsequent progression. In contrast, p53 abnormalities were present in 57/87 (65.5%) of these endoscopies (Chi square P<0.00001 for both), **Figure 3C-D**. Most pre-progression biopsies with a morphologic abnormality were also p53-ABNL with only 11/231 (4.8%) being p53-NL. Overall, p53-ABNL was much more prevalent in pre-progression biopsies, occurring more frequently and earlier, than any morphologic abnormality (BE-IND and BE-LGD combined).

**Figure 3.**
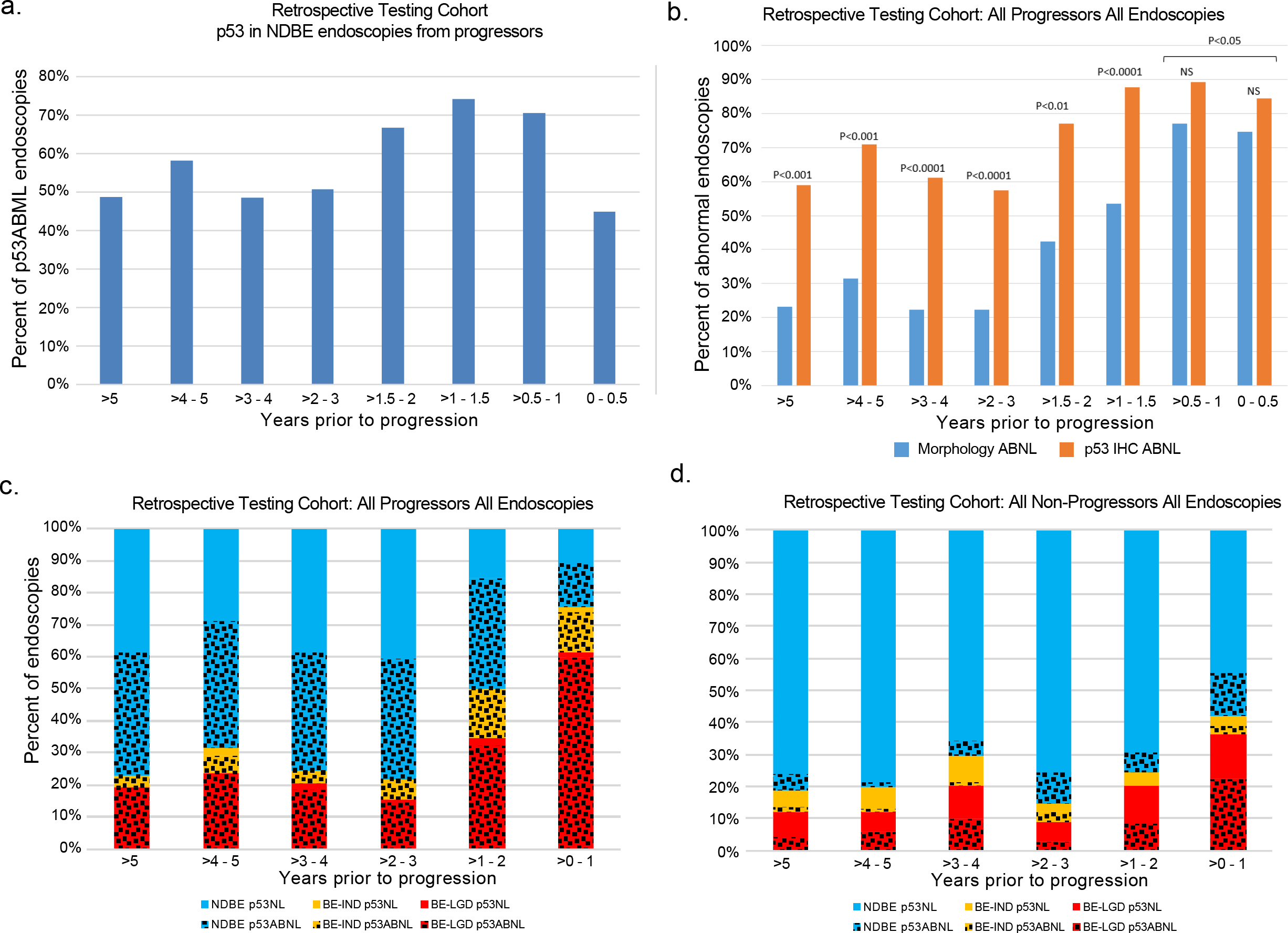
p53-ABNL vs time to progression. a) Percent of endoscopies from patients in the baseline NDBE cohort that were p53-ABNL taken at different time points before progression. b) Comparison of the percentage of endoscopies positive for either a histologic abnormality (BE-IND or BE-LGD) or p53 IHC broken down over different time periods before progression. c-d) Frequency of first morphologic or first p53 abnormality in all progressors (c) or non-progressors (d). At every point in time, more than 50% of progressors have had at least one p53-ABNL biopsy, while substantially less than 50% have had a morphologic abnormality until the final pre-progression year.

### P53 IHC as biomarker for progression

**Table 2** summarizes the sensitivities and specifies for progression of p53-ABNL in the different histologic diagnoses at either the baseline endoscopy or across all endoscopies. In our NDBE case-control testing cohort, p53-ABNL in the baseline endoscopy had a sensitivity of 49.7% and specificity of 98.3% for progression with an odds ratio of 58 (95% CI 17.9-188.5, P<0.0001). In this case-control study, there were 127 patients with progression where all endoscopies (baseline + surveillance) before progression had exclusively NDBE and there was greater than 1 year between baseline endoscopy and progression. In this group of subjects, who by all current clinical standards were considered low risk throughout their surveillance before progression, 72/127 (56.7%) had a p53-ABNL endoscopy before progression leading to a sensitivity of 56.7% and specificity of 92.2%. The odds ratio of progression for p53 positivity in patients with exclusively NDBE diagnosis and with biopsies taken at least 1 year prior to progression was 15.4 (95% CI 8.1-29.5, P<0.0001). We next limited analysis to only endoscopies performed at least 5 years before progression or end of follow up in NDBE patients to determine how p53 IHC performed at or outside the current recommended surveillance interval. The sensitivity and specificity of p53-ABNL for progression was still 46.2% and 96.4% respectively, supporting potential longer follow up intervals in p53-NL NDBE.

**Table 2.** Sensitivity and specificity of p53 IHC for detection of progression.

| **Diagnosis** | **Sensitivity** | **Specificity** | **Odds Ratio** |
| --- | --- | --- | --- |
| NDBE at baseline only | 49.7% | 98.3% | 58.0 |
| NDBE prior to any BE-IND/BE-LGD | 57.5% | 93.3% | 18.9 |
| BE-IND at baseline only | 90.0% | 84.6% | 49.5 |
| BE-IND prior to any BE-LGD | 92.3% | 69.2% | 27.0 |
| BE-LGD at baseline only | 94.2% | 54.6% | 17.8 |
| BE-LGD prior to any BE-HGD/BE-CA | 93.9% | 43.2% | 11.6 |

Results in this analysis are summarized for each endoscopy (ie. All biopsies at a given time-point)

In our abnormal histology testing cohort, p53-ABNL in the baseline endoscopy with a diagnosis of IND was associated with a sensitivity of 90.0% and specificity of 84.6% for progression with an odds ratio of 49.5 (95% CI 10.0-245.0, P<0.0001). p53-ABNL in the baseline endoscopy with a diagnosis of LGD was associated with sensitivity of 94.2% and specificity of 54.6% for progression with an odds ratio of 17.8 (95% CI 6.4-49.5, P<0.0001). If the control group was restricted to only patients who had two or more endoscopies with confirmed LGD, the results held with a slightly increased specificity of 66.7%.

As examples where a biomarker in NDBE samples would have been highly valuable clinically, in our study set there were 32 subjects with NDBE who progressed to fully invasive esophageal adenocarcinoma while on surveillance. Histologic abnormalities were identified prior to invasive carcinoma progression in only 6/32 (18.8%), **Figure S4**. Among the remaining 26 individuals, histology failed to alert the clinicians these patients were at high risk of developing cancer. However, abnormal p53 IHC was present in all six subjects with histologic abnormalities, and in an additional 11/26 (42.3%) subjects who only had NDBE diagnosed before developing invasive EAC. Overall, p53 IHC was significantly more sensitive than histology at identifying an abnormality prior to the development of invasive EAC (6/32 vs 17/32, Fisher exact P<0.01). Among these 32 cases, there were 24 subjects who progressed from NDBE to invasive cancer before their next recommended endoscopy (interval progression). Regardless of whether a higher-grade lesion was not biopsied in the prior screening endoscopy or these patients rapidly progressed, a biomarker warning the clinicians that these patients were at heightened risk and should be followed more closely would likely have enabled earlier diagnosis or prevention. In these 24 subjects, 11 (46%) were p53-ABNL.

### p53 IHC stratifies patients in a routine practice setting

We next wanted to determine whether the addition of p53 IHC to standard histologic diagnosis would improve Barrett’s outcome prediction in a routine clinical setting. Prospective p53 IHC, using the same scoring criteria described above, was adopted for routine signout of clinical BE biopsies at Inform Diagnostics in June 2011. Pathology records were searched to identify all BE patients with a reported p53 IHC result and a subsequent follow-up biopsy. Of 389 cases of BE-IND and 389 cases of BE-LGD identified with p53 IHC, 22 (5.6%) and 78 (20%) progressed to either HGD or EAC, respectively. Amongst 650 NDBE cases, in the limited follow-up time, 20 (3.1%) progressed to LGD and 10 (1.5%) progressed to HGD/EAC. Kaplan-Meier analysis (**Figure 4**) showed the ability of p53 IHC to clearly stratify patients in all cases (Log-rank test, z = 3.81 P<0.001 for BE-IND, z = 3.76 P<0.001 for BE-LGD, z = 9.81 P<0.001 for NDBE to LGD, and z = 3.08 P=0.002 for NDBE to HGD). In total, p53 IHC was able to stratify patients with respect to progression regardless of the histologic diagnosis in a large prospective cohort in a standard clinical setting.

**Figure 4:**
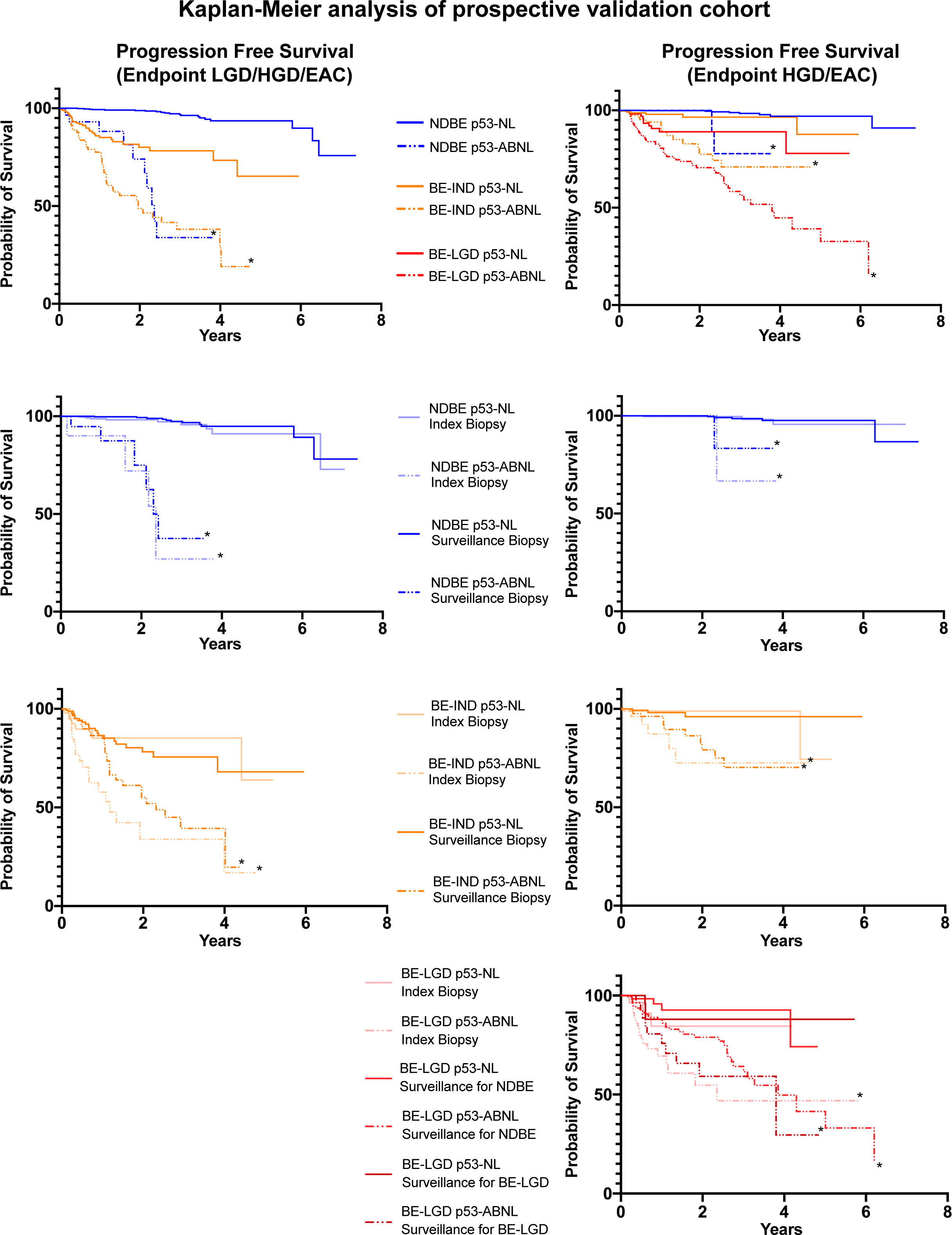
p53 IHC patient stratification in a prospective validation cohort. Kaplan-Meier curves for progression to either BE-LGD/BE-HGD/EAC (left side) or to BE-HGD/EAC (right side) free survival in patients with p53-ABNL and p53-NL. All patient numbers and statistical analysis is reported in supplementary table S10. * denotes p<0.05 for p53-NL vs p53-ABNL.

To determine if the timing of the biopsy in relation to the patient’s surveillance history may affect the results, we separated the results into the patients with p53 IHC information on their first index endoscopy (first ever diagnosis of Barrett’s esophagus), patients where the first p53 IHC was on a surveillance endoscopy for NDBE (no known BE-IND or BE-LGD), or BE-LGD patients where the first p53 IHC was on a surveillance endoscopy following a diagnosis of BE-LGD. Kaplan-Meier curves for progression free survival were similar for patients with an index sample or patients with only surveillance endoscopies in NDBE and BE-IND. For BE-LGD, patients identified in an index endoscopy did slightly worse than those undergoing surveillance for NDBE (log-rank test; z=2.94, p=0.014), **Supplementary Figure S8**. Under all conditions of having the index endoscopy or only surveillance endoscopies and under all histologic diagnoses, p53 IHC was able to stratify progressors from non-progressors, confirming the results seen in the retrospective testing cohorts (**Figure 4** and **Table S10)**.

## Discussion

EAC and its associated precursor, BE, is a growing clinical problem. While it is recognized that BE predates most EAC, and screening and surveillance paradigms have been established, we still fail to identify the majority of cancer patients early when they are most treatable. In addition to the many people not getting the treatment they need (underdiagnosis), there is a large population of people being overtreated (overdiagnosis) due to the very low rate of NDBE progression and the growing use of endoscopic treatment for cases of BE-LGD (with a widely variable though universally fairly low rate of progression). Thus, there is a clear need for improved methods and biomarkers for risk stratification of patients with BE. Many studies have focused on the different molecular alterations present in BE, and the best risk stratification will likely come from a combination of molecular biomarkers^11,12,14,19,26–30^. However, several barriers still exist before widespread clinical adaptation of such testing can be accomplished. Here we present data to show that an inexpensive readily available biomarker could have immediate strong clinical utility. Our data greatly expands and compliments previous studies examining p53 and Barrett’s esophagus. In one of the most cited studies, Kastelein et. al. looked at p53 IHC for risk prediction and used grading criteria similar to ours. In their well-controlled, case-control study they had 15 patients with NDBE and 34 patients with LGD who progressed to either HGD or EAC and similar to our study found that 32.4% and 70.7% of biopsies from NDBE and LGD progression patients respectively had abnormal p53 IHC^18^. While this study utilized strictly controlled biopsy sampling including 4 quadrant biopsies every 2 cm of BE and performed all p53 IHC staining as a single batch, for a biomarker to be clinically useful it needs to function under more ‘real world’ settings which unfortunately often fail to meet these high standards.^31^ Our study utilized samples taken in the community setting under routine care and with a substantially higher number of progression patients. In the prospective validation cohort, this included in real-time IHC staining from 3 different laboratories and interpretation from over 40 different pathologists. In light of past studies, we feel that obtaining our results under these ‘real world’ conditions strongly support the robustness and reproducibility of using p53 IHC.

In the setting where the risk of progression is low, such as for subjects with NDBE, a useful biomarker will require a high specificity in order to reduce the number of false positives inherent to this situation. In our cohort of patients with only NDBE prior to progression, p53-IHC was quite specific. The sensitivity was lower, but still identified half of all progressors in a cohort of patients who showed no histologic abnormalities (i.e. no BE-IND or BE-LGD) before progression. If one assumes a general NDBE progression rate of 0.3% per year, our data would suggest an approximately 30% risk of progression within 5 years for patients with p53-ABNL NDBE. While the estimates for progression risk in LGD is highly variable, this calculated rate for risk of progression p53-ABNL NDBE is comparable to rates of progression in those diagnosed with BE-LGD. Given this, changing practice to treat p53-ABNL NDBE as an equivalent to how we now treat BE-LGD with either yearly endoscopies or consideration of ablation would seem appropriate. Current guidelines suggest endoscopic surveillance every 3-5 years for non-dysplastic Barrett’s esophagus, with trends leaning toward closer to the 3 years as opposed to 5 years. While not guideline changing, it would seem quite reasonable to lean toward the 5-year mark for p53 normal NDBE cases and thus reducing overall number of endoscopies. With the low cost and ease of implementation, p53 IHC could be a valuable addition to standard histologic workup of screening and surveillance NDBE biopsies.

Currently, many patients with BE-LGD are undergoing treatment to eradicate their BE. While potentially helpful for those with BE-LGD that will progress to cancer, studies suggest the majority of these patients would not progress if left untreated^32–36^. With endoscopic treatment for BE-LGD becoming standard, for a biomarker to be useful it needs to be highly sensitive to not miss patients where treatment would be beneficial but allow the identification of some lower risk patients where alternative approaches besides treatment may be better suited. In BE-LGD patients, p53 IHC identified those who would progress with a high sensitivity of 94%. While a proportion of the non-progression subjects were also positive, p53 IHC still provided increased stratification and identified a low risk group in this difficult to manage population while identifying the vast majority of patients who will progress. Given p53’s high sensitivity, a strong argument can be made to spare BE-LGD p53-NL patients from the potential morbidity/complications of therapy, whereas a p53-ABNL result would lead to a firm recommendation to receive ablation. Several studies have also shown a high inter-observer variability in pathologists diagnosing BE-LGD, adding further difficulties to the clinical decision making process^6,7^. A biomarker that can identify the vast majority of patients who will progress may reassure clinicians and patients that their decision for continued surveillance or therapy is sound.

The diagnosis of BE-IND is challenging to manage and is considered to be a mixture of patients with NDBE that has developed intense reactive changes and patients who have a true dysplasia. In our prospective cohort, the p53-ABNL BE-IND patients progressed similarly to patients with a BE-LGD diagnosis. As both progress at nearly equal rates, clinically it would not be unreasonable to treat patients with p53-ABNL BE-IND in a similar manner as BE-LGD. The p53-NL BE-IND patients had a significantly lower progression rate with only 7 of 263 progressing through the duration of the study. Given this low rate of progression, these patients could likely undergo surveillance less frequently.

The potential utility of p53 IHC as a biomarker of Barrett’s progression risk has been studied in the past. While these studies showed promise, many of them have either looked at its use exclusively in the setting of dysplasia, had limited numbers of NDBE progressors that hindered broad conclusions to be drawn, failed to account for the p53-absent staining pattern, and/or had other limitations that deterred widespread adoption^15–18,26,37,38^. Our study removes these barriers by including a large number of NDBE progression patients (179), having samples from community-based practices throughout the United States, having grading criteria designed to maximize specificity while preserving sensitivity, and having a prospective validation cohort. Given the previously reported results and our findings, there is clear evidence to support p53 IHC use as an adjuvant to routine histologic analysis.

## Data Availability

All data within the manuscript is available either within the manuscript or through communication with the authors.

## METHODS

### p53 immunohistochemistry

Immunohistochemical stains for p53 were performed using the DO-7 antibody on the BenchMark XT or BenchMark ULTRA automated slide staining systems with the OptiView or UltraView detection kits (Ventana Medical Systems, Tucson, AZ) according to the manufacturer’s recommendations. Positive and negative controls were included in all staining runs. The percentage of nuclei with positive staining was scored on an intensity scale of 0-3, with 0+ representing no staining and 3+ representing very strong staining (**Figure 1B**). Abnormal (p53-ABNL) staining was considered either 2-3+ nuclear positivity in >50% of cells in at least one crypt base or glandular profile, or within a contiguous focus of at least 20 surface epithelial cells. Alternatively, p53-ABNL was also identified in case of total absence of staining (0+) in all epithelial cells of at least one crypt base or glandular profile, or strong cytoplasmic staining with complete absence of nuclear staining (rare).

### Development of p53 IHC scoring criteria

While much has been published regarding abnormal p53 expression in cancers and dysplasia, much less is known about expression patterns in NDBE. To further delineate the spectrum of p53 IHC staining in non-dysplastic biopsies, and delineate criteria for abnormal staining, we performed p53 IHC in 18 NDBE biopsies from patients with no known dysplasia, and in 115 NDBE biopsies that were concurrent with high grade dysplasia. The percentage of nuclei with positive staining was scored on an intensity scale of 0-3, with 0+ representing no staining and 3+ representing very strong staining. A consistent pattern of crypt staining was seen in all crypts in NDBE biopsies from patients with no known dysplasia, and in most crypts in NDBE biopsies from patients with concurrent high-grade dysplasia: crypt base positivity was always present, with most cells having 0-1+ nuclear positivity, scattered cells having 2+ nuclear positivity, and rare cells having 3+ nuclear positivity (Figure S5). This crypt base positivity diminished towards the surface, which typically had 0 to scattered 1+ IHC positivity. This staining pattern is consistent with the physiologic normal expression of p53 in the proliferative zone of the crypt base.

In addition to this normal p53 crypt expression pattern, NDBE biopsies from patients with concurrent high-grade dysplasia harbored foci of distinctly abnormal staining comprised of two common patterns: markedly increased staining, and complete absence of staining (**Supplementary Figure S5**). Increased p53 expression is known to be associated with *TP53* point mutations; these result in protein stabilization and nuclear accumulation. Absent p53 expression is known to be associated with *TP53* truncation mutations, and other genomic aberrations that result in loss of p53 expression. To develop criteria for abnormally increased staining, we identified the foci that had the strongest staining, and scored the percentage of cells that had either 2+ and 3+ nuclear positivity. Increased staining extended onto the luminal surface in some cases, but was confined to the crypt base in others, in which case only the crypt base was evaluated. Foci as small as a single individual crypt base were considered adequate for scoring increased staining. Scoring using either 3+ positivity alone, or combined 2-3+ positivity, revealed that approximately 35% of NDBE biopsies from patients with concurrent high-grade dysplasia had foci of increased p53 expression that was greater than anything present in the NDBE biopsies without dysplasia (**Supplementary Figure S6**). We chose >50% 2-3+ nuclear positivity as a cut-off to define abnormal p53 expression because it had 100% specificity for an association with concurrent high-grade dysplasia, and could be rapidly assessed by a pathologist, potentially making it a practical biomarker for routine clinical use. Finally, occasional NDBE biopsies from patients with concurrent high-grade dysplasia also had focal marked p53 positivity present only within surface epithelium, but not adjacent crypts. This finding was consistent with a mutant p53 clone extending onto the surface from a nearby crypt not present in the section. Therefore, the presence of 2-3+ p53 positivity in >50% of a contiguous focus of 20 surface cells was also considered abnormal.

While the vast majority of crypts from NDBE biopsies with no known dysplasia had ≤20% 2-3+ nuclear positivity in crypt bases, there were some crypts with 31-40% 2-3+ nuclear positivity. Although this degree of positivity was present at higher frequency in NDBE biopsies from patients with concurrent high-grade dysplasia, there was appreciable overlap. Therefore, 2-3+ nuclear positivity in 21-50% of crypt epithelial nuclei was considered equivocal, and while underlying *TP53* mutations may be present in a subset of these cases, was classified as p53-NL in all analyses in this study.

The second common abnormal p53 expression pattern, complete absence of p53 expression, was identified in 20/115 (17%) NDBE biopsies from patients with concurrent high-grade dysplasia compared to none of the NDBE biopsies from patients with no known dysplasia. Because of the consistent normal p53 expression pattern present in crypt bases of NDBE biopsies from patients with no known dysplasia, this abnormal loss of staining could be reliably scored when identified in foci as small as a single crypt base. In contrast to abnormally increased p53 expression, the very low (normal) p53 expression in Barrett’s epithelium at the luminal surface makes it impossible to accurately diagnose absent p53 expression (mutation) when the involvement is limited to the surface epithelium of a NDBE biopsy.

The criteria defined above were further validated in an additional sample set of 50 NDBE biopsies from patients with no known dysplasia, and 50 BE-HGD biopsies. Abnormal p53 expression was found in 2/50 (4%) of the NDBE biopsies and 48/50 (96%) BE-HGD biopsies, providing additional validation that these scoring criteria are very sensitive for identification of abnormal p53 in advanced Barrett’s neoplasms, while only being positive in a very small subset of unselected NDBE biopsies (which is to be expected for a possible biomarker).

Finally, during the blinded slide review of the retrospective cohort, we identified one additional distinctive rare abnormal p53 expression pattern: complete absence of nuclear p53 positivity in the presence of strong cytoplasmic positivity. This abnormal pattern has been seen for some other nuclear proteins as well as in p53 and has been attributed to mutations that result in loss of the nuclear localization domain^39^. This abnormal pattern was considered to be a variant of loss of expression and was combined with absent staining pattern in subsequent analyses. **Supplementary Table S1** summarizes the p53 expression criteria used in this study.

### TP53 sequencing

In samples that had at least one remaining unstained slide, areas of BE were macrodissected for DNA isolation and sequencing of all exons of the *TP53* gene. Library construction was performed on any sample with at least 2ng of total DNA using the Pillar Biosciences (Natick, MA) SLIMAmp target enrichment technology, whereby all exons of *TP53* were amplified with multiplexed PCR and the resulting material utilized for next-generation sequencing. Samples were pooled and sequenced on an Illumina Miseq. The Pillar PIVAT analysis software was used to analyze the results.

To determine likely somatic, pathogenic *TP53* mutations, as series of filtering steps were performed. All mutational calls under 0.025 allele frequency, present in any ethnic background at > 0.001 in the gnomAD database (https://gnomad.broadinstitute.org/about), or were not within the coding region or splice region were discarded. Additionally, missense mutations that have not been previously reported in esophageal adenocarcinoma (cbioPortal) or were not reported in the COSMIC database at least 5 times were removed. Predicted nonsense, frameshift, or splice altering mutations were retained.

### Patient Selection (retrospective testing cohort)

To identify a diverse, clinically relevant collection of BE samples a search of Inform Diagnostics pathology records was performed and identified 313 potential non-progressors and 359 potential progressors, for a total of 672 eligible study subjects (**Supplementary Table S2**). Pathology samples in the records consisted of patients from predominately community-based practices throughout the United States that utilize Inform Diagnostics for their pathology services. There were 111 subjects (16.5%) excluded because of lack of availability of slides or tissue, or discrepant central pathology review that excluded the subject from further study (**Supplementary Table S3**). A total of 46 subjects (6.8%) were re-classified from one study group to another, predominantly because of a change in baseline diagnosis during central pathology review (**Supplementary Table S4**). After exclusions and re-classifications, a total of 249 non-progressors and 312 progressors were included in the study. To look at identification of both possible incident BE-HGD or OAC and subsequent progression and to obtain a complete picture of the timing of acquisition of p53 abnormalities prior to progression, all available samples/time points before progression were included. Baseline endoscopies were defined as the first endoscopy with a diagnosis of Barrett’s esophagus seen at Inform diagnostics. Clinical characteristics of subjects are summarized in **Table 1**. The characteristics between cases and controls were comparable. However, baseline BE-IND and BE-LGD progressors were less likely to be female than non-progressors. There was no association detected between age at diagnosis and p53 IHC status (Table S5). We chose to study BE-IND and BE-LGD cases as these patients can be clinically challenging to manage since there is significant inter/intra pathologist disagreement for these diagnoses, leading to widely variable estimations of progression risk. We included cases who may have had incident progression (defined as NDBE, BE-IND, BE-LGD samples diagnosed within 1 year of a BE-HGD or OAC diagnosis) as even with modern endoscopic techniques, focal HGD and early OAC may be missed during endoscopic screening.

### Patient Selection (prospective validation cohort)

Starting in 2011, Inform diagnostics began performing p53 IHC as part of routine Barrett’s pathology work up. p53 IHC was performed in three different laboratories with one each in Massachusetts, Texas, and Arizona. A total of 41 pathologists signed out the p53 IHC results during their clinical review of the case. Patients were divided by histologic diagnosis of their baseline endoscopy as well as whether the baseline diagnosis was the patients true index endoscopy or whether they were in a surveillance program for NDBE or LGD. All patients with prior endoscopic/surgical therapy were not included in the study. For NDBE and BE-IND, only patients undergoing surveillance for NDBE (ie no history of BE-IND or BE-LGD) were included in the surveillance baseline group.

### Central Pathology Review

For the retrospective testing cohorts (**Figure 1**), to ensure accurate and consistent pathologic diagnoses of the specific samples used for the above analyses, when tissue blocks of archival BE samples were processed to generate slides for p53 IHC, new hematoxylin and eosin (H&E) stained slides were created and submitted for pathologic review. Pathologists were blinded to outcome and p53 status. When there were discrepancies between the original diagnosis and reviewed diagnosis, a third independent reviewer assessed the slides. Any remaining discrepancies were assessed by consensus at a multi-headed microscope. Primary analysis was performed using the central review pathologic diagnosis. A secondary analysis using the original pathologic diagnosis was also performed to validate the utility of p53 IHC in the context of the original routine clinical diagnosis. For the prospective cohort, to determine how the p53 IHC protocol would work in routine practice, we utilized the original clinical diagnosis except for all diagnoses of dysplasia or cancer were confirmed by at least one additional pathologist.

For the retrospective testing cohorts, central pathology review confirmed the presence of Barrett’s mucosa, columnar dysplasia or adenocarcinoma in 3243 biopsies. IHC for p53 was not attempted in 353 of these cases because there was no tissue available, or the histologic focus of interest was not present in remaining tissue. Of the remaining 2890 biopsies, a p53 IHC result was obtained in 2853 (98.7%). **Figure 1** shows examples of histologic findings and p53 IHC staining from both non-progressors and progressors, across the complete spectrum of precursor diagnoses in this study.

### Statistical analysis

A Fisher’s exact test was used to compute P values from a 2 × 2 contingency tables. A t-test was used to compare mean values of two (continuous variable) groups. A log rank test was used to compare Kaplan-Meier curves between p53-ABNL and p53-NL groups. All P values were two-tailed.

