## Supplementary tables and figures for "Abnormal *TP53* predicts risk of progression in patients with Barrett’s esophagus regardless of a diagnosis of dysplasia"

**Supplementary Appendix**

| | |
|---|---|
| Supplementary Figure Legends | pg 2 |
| Supplementary Tables | pg 3 |
| Supplementary Figures | pg 13 |

**Supplementary Figure Legends**

**Supplementary Figure S1. Cartoon of endoscopy and biopsy terminology.** Diagram illustrates determination of baseline or later surveillance endoscopies as positive (p53-ABNL) or negative (p53-NL) by biopsies. When looking at the endoscopy level, if any biopsy/pathology block at a given endoscopy is p53-ABNL the endoscopy is considered to be positive/abnormal.

**Supplementary Figure S2.** Accession timeline chart of baseline NDBE p53-NL progressors with follow-up NDBE biopsies prior to development of dysplasia. For the progression patients, multiple cases had a p53-NL baseline endoscopy but a subsequent p53-ABNL endoscopy (BE-CA samples diagnosed with esophageal adenocarcinoma).

**Supplementary Figure S3. p53-ABNL vs time to progression.** The percentage of p53-ABNL endoscopies for progression patients was plotted as a function of time for each histologic diagnosis.

**Supplementary Figure S4. Timeline charts of subjects with intramucosal (A) and invasive carcinoma (B).** Multiple patients had progression from NDBE to invasive cancer within the recommended surveillance interval without a diagnosis of dysplasia. The timing of the different endoscopies, p53 status, and histologic diagnosis are plotted.

**Supplementary Figure S5. p53 IHC scoring** A) example of cells negative p53 IHC (blue arrow), 1+ staining (tan arrow), 2+ staining (light brown arrow), and 3+ staining (dark brown arrow). B) example of wild type staining. C) Example of positive nuclear staining (p53-ABNL). D) Example of absent staining (p53-ABNL). Glands quantified in Blue circles.

**Supplementary Figure S6. Distribution of p53 IHC positivity in NDBE biopsies.** Bar chart showing numbers of samples with grading of 0, 1+, 2+, 3+ p53 IHC in NDBE biopsies.

**Supplementary Figure S7. Kaplan-Meier analysis of progression free survival in patients with screening or surveillance baseline endoscopies.** Kaplan-Meier analysis for progression free survival split between patients with a true index endoscopy as baseline or a surveillance endoscopy for NDBE or LGD as baseline. Baseline is defined as the first endoscopy with a diagnosis of Barrett's esophagus seen at Inform Diagnostics. Progression from NDBE or BE-IND to LGD, HGD, or OAC (top) and NDBE, BE-IND, or LGD to HGD or OAC (bottom).

### SUPPLEMENTARY TABLES

**Supplementary Table S1. p53 immunohistochemistry scoring criteria**

| **p53 Expression Pattern** | **Criteria** | **Categorization for Analyses** |
|---|---|---|
| Normal (wild-type) pattern | 2-3+ nuclear positivity in $\leq$20% of epithelial cells in all individual crypts or glandular profiles, and all contiguous stretches of surface epithelium | p53-NL |
| Equivocal pattern | 2-3+ nuclear positivity in 21-50% of cells in at least one crypt base or glandular profile, or within a contiguous focus of at least 20 surface epithelial cells | p53-NL |
| Point mutation pattern | 2-3+ nuclear positivity in >50% of cells in at least one crypt base or glandular profile, or within a contiguous focus of at least 20 surface epithelial cells | p53-ABNL |
| Absent (null) pattern | Total absence of staining in all epithelial cells of at least one crypt base or glandular profile* | p53-ABNL |
| Cytoplasmic pattern | Absence of nuclear staining with aberrant cytoplasmic staining in all epithelial cells of at least one crypt base or glandular profile* | p53-ABNL |

*Diagnosing the absent or cytoplasmic patterns required the presence of internal control positivity (nuclear positivity in crypt epithelium, squamous epithelium, immune cells or stromal cells) as evidence of stain adequacy.

**Supplementary Table S2. Study subject eligibility and inclusion**

| | Eligible | Excluded (%)[1] | Included same class (%) | Included Re-classified (%)[2] | Final Total Subjects (with re-classified) |
|---|---|---|---|---|---|
| Total subjects | 672 | 111 (16.5%) | 515 (76.6%) | 46 (6.8%) | 561 |
| Non-progressors | 313 | 63 (20.1%) | 241 (77.0%) | 9 (2.9%) | 249 |
| NDBE | 195 | 16 (8.2%) | 179 (91.8%) | 0 | 179 |
| BE-IND | 53 | 27 (50.9%) | 22 (41.5%) | 4 (7.5%) | 26 |
| BE-LGD | 65 | 20 (30.8%) | 40 (61.5%) | 5 (7.7%) | 44 |
| Progressors | 359 | 48 (13.4%) | 274 (76.3%) | 37 (10.3%) | 312 |
| NDBE | 196 | 10 (5.1%) | 170 (86.7%) | 16 (8.2%) | 179 |
| BE-IND | 45 | 9 (20%) | 22 (48.9%) | 14 (31.1%) | 30 |
| BE-LGD | 118 | 29 (24.6%) | 82 (69.5%) | 7 (5.9%) | 103 |

[1]See Supplementary Table 1 for details of excluded cases.

[2]Most re-classifications were due to a central path review change of the baseline diagnosis; see Supplementary Table 4 for details.

**Supplementary Table S3. Details of excluded study subjects**

| Category | Total | Details |
|---|---|---|
| Non-progressors | 63 | |
| NDBE | 16 | Baseline not available: 15 |
| | | Final follow-up biopsy upgraded to BMID: 1 |
| BE-IND | 27 | Baseline downgraded to BM: 22 |
| | | Baseline not available: 4 |
| | | Final follow-up upgraded to BMLD: 1 |
| BE-LGD | 20 | Baseline downgraded to BM: 14 |
| | | Baseline not available: 5 |
| | | Baseline upgraded to BMHD: 1 |
| Progressors | 48 | |
| NDBE | 10 | Baseline not available: 6 |
| | | Progression not confirmed: 4 |
| BE-IND | 9 | Baseline not available: 4 |
| | | Baseline upgraded to BMHD: 4 |
| | | Progression not confirmed: 1 |
| BE-LGD | 29 | Baseline upgraded to BMHD: 19 |
| | | Baseline NA: 9 |
| | | Progression not confirmed: 2 |

**Supplementary Table S4. Details of re-classified study subjects**

| Category | | Total | Details |
|---|---|---|---|
| Non-progressor to non-progressor | | 8 | |
| | BE-IND to BE-LGD | 4 | Baseline upgraded from BE-IND to BE-LGD |
| | BE-LGD to BE-IND | 4 | Baseline downgraded from BE-LGD to BE-IND |
| Non-progressor to progressor | | | |
| | BE-LGD to NDBE | 1 | Final follow-up upgraded from BE-LDG to BE-HGD; NDBE prior to first BE-LGD available for new baseline |
| Progressor to progressor | | 37 | |
| | NDBE to BE-IND | 4 | Baseline upgraded from NDBE to BE-IND: 3<br>Baseline not available: 1 |
| | NDBE to BE-LGD | 12 | Baseline upgraded from NDBE to BE-LGD: 8<br>Baseline not available: 4 |
| | BE-IND to NDBE | 5 | Baseline downgraded from BE-IND to NDBE: 5 |
| | BE-IND to BE-LGD | 9 | Baseline upgraded from BE-IND to BE-LGD: 7<br>Baseline not available: 2 |
| | BE-IND to NDBE | 3 | Baseline downgraded from BE-IND to NDBE: 3 |
| | BE-LGD to BE-IND | 4 | Baseline downgraded from BE-LGD to BE-IND: 4 |
| Progressor to non-progressor | | 0 | Progression not confirmed: 7, all excluded from study |

**Supplementary Table S5. p53 IHC vs age**

| | Total | Youngest age quartile | Lower middle age quartile | Upper middle age quartile | Oldest age quartile | |
|---|---|---|---|---|---|---|
| *All non-progressors* | | | | | | |
| NDBE | 943 | 9/236 (3.8%) | 13/236 (5.5%) | 21/236 (8.9%) | 15/235 (6.4%) | NS |
| BE-IND | 63 | 2/16 (12.5%) | 3/16 (18.7%) | 2/16 (12.5%) | 2/15 (13.3%) | NS |
| BE-LGD | 121 | 14/31 (46.7%) | 10/30 (33.3%) | 12/30 (40.0%) | 5/30 (16.7%) | P<0.05 for oldest quartile vs all others |
| *All Progressors* | | | | | | |
| NDBE | 620 | 68/155 (43.9%) | 62/155 (40.0%) | 73/155 (47.1%) | 66/155 (42.6%) | NS |
| BE-IND | 76 | 16/19 (84.2%) | 17/19 (89.5%) | 17/19 (89.5%) | 19/19 (100%) | NS |
| BE-LGD | 298 | 69/75 (92.0%) | 68/75 (90.7%) | 73/74 (98.6%) | 72/74 (97.3%) | NS |

**Supplementary Table S6. p53 IHC abnormalities in baseline endoscopies summarized for each subject[1]**

| Diagnostic category at baseline | Progression Status | Total patients with specified baseline diagnosis | Number of specified baseline endoscopies with abnormal p53 IHC | Percent | Chi square P-value[1] |
|---|---|---|---|---|---|
| NDBE | Non-progressor | 179 | 3 | 1.7% | |
| | Progressor | 179 | 89 | 49.7% | <0.00001 |
| BE-IND | Non-progressor | 26 | 4 | 15.4% | |
| | Progressor | 30 | 27 | 90.0% | <0.00001 |
| BE-LGD | Non-progressor | 44 | 20 | 45.4% | |
| | Progressor | 103 | 97 | 94.2% | <0.00001 |

[1]The results of baseline biopsies in individual patients were summarized as follows: if there were multiple biopsies with Barrett's mucosa negative for dysplasia, the subject was considered to have abnormal p53 IHC if any one biopsy was abnormal.

[2]Non-progressor vs progressor for each diagnostic category

**Supplementary Table S7. p53 IHC abnormalities at baseline with each block/biopsy counted separately**

| Diagnostic category at baseline | Progression Status | Total pathology blocks with specified baseline diagnosis | Number of specified baseline pathology blocks with abnormal p53 IHC | Percent | Chi square P-value[1] |
|---|---|---|---|---|---|
| NDBE | Non-progressor | 219 | 3 | 1.4% | |
| | Progressor | 279 | 112 | 40.1% | <0.00001 |
| BE-IND | Non-progressor | 29 | 3 | 10.3% | |
| | Progressor | 32 | 27 | 84.8% | <0.00001 |
| BE-LGD | Non-progressor | 55 | 20 | 36.4% | |
| | Progressor | 131 | 122 | 93.1% | <0.00001 |

[1]Non-progressor vs progressor for each diagnostic category

**Supplementary Table S8. p53 IHC abnormalities in baseline biopsies/pathology blocks using original final diagnosis (not central pathology review results)**

| Diagnostic category at baseline | Total biopsies with specified baseline diagnosis | Number of specified baseline biopsies with abnormal p53 IHC | Percent | Chi square P-value[1] |
|---|---|---|---|---|
| *Non-progressor* | | | | |
| Negative for dysplasia | 194 | 3 | 1.5% | |
| Indefinite for dysplasia | 59 | 10 | 15.1% | |
| Low grade dysplasia | 71 | 26 | 36.6% | |
| *Progressor* | | | | |
| Negative for dysplasia | 259 | 106 | 40.9% | <0.00001 |
| Indefinite for dysplasia | 38 | 26 | 68.4% | <0.00001 |
| Low grade dysplasia | 139 | 129 | 92.8% | <0.00001 |

[1]Non-progressor vs progressor

**Supplementary Table S9. p53 IHC abnormalities in baseline endoscopies summarized for each subject using original final diagnosis (not central pathology review results)**

| Diagnostic category at baseline | Total subjects in baseline diagnostic category | Number of subjects with abnormal p53 IHC in any baseline biopsy | Percent | Chi square P-value[1] |
|---|---|---|---|---|
| *Non-progressor* | | | | |
| Negative for dysplasia | 179 | 3 | 1.7% | |
| Indefinite for dysplasia | 27 | 5 | 18.5% | |
| Low grade dysplasia | 38 | 18 | 47.4% | |
| *Progressor* | | | | |
| Negative for dysplasia | 179 | 89 | 51.1% | <0.00001 |
| Indefinite for dysplasia | 33 | 24 | 72.7% | <0.0001 |
| Low grade dysplasia | 96 | 90 | 93.7% | <0.00001 |

[1]Non-progressor vs progressor

**Table S10. p53 IHC abnormalities in Prospectively tested validation cohort separated by index and surveillance endoscopies**

| Patient Category | Diagnosis of Index p53 Biopsy | p53 IHC Normal | p53 IHC Abnormal | Progression Free Survival Endpoint | Log Rank Test | P-Value |
|---|---|---|---|---|---|---|
| p53 IHC on first Barrett's biopsy (Index) | | | | | | |
| | NDBE | 260 | 10 | LGD/HGD/CA | Z=6.52 | P<0.001 |
| | | | | HGD/CA | Z=2.81 | P=0.0049 |
| | BE-IND | 97 | 41 | LGD/HGD/CA | Z=3.99 | P<0.001 |
| | | | | HGD/CA | Z=2.09 | P=0.037 |
| | BE-LGD | 30 | 79 | HGD/CA | Z=1.97 | P=0.049 |
| p53 IHC on known Barrett's surveillance biopsy (no prior BE-IND or dysplasia) | | | | | | |
| | NDBE | 356 | 20 | LGD/HGD/CA | Z=10.07 | P<0.001 |
| | | | | HGD/CA | Z=2.65 | P=0.0081 |
| | BE-IND | 161 | 90 | LGD/HGD/CA | Z=2.98 | P=0.0029 |
| | | | | HGD/CA | Z=2.19 | P=0.0014 |
| | BE-LGD | 65 | 174 | HGD/CA | Z=2.61 | P=0.0092 |
| p53 IHC on known BE-LGD patient | | | | | | |
| | BE-LGD | 26 | 30 | HGD/CA | Z=2.08 | P=0.038 |

**SUPPLEMENTARY FIGURES**

**Supplementary Figure S1. Cartoon of endoscopy and biopsy terminology.**

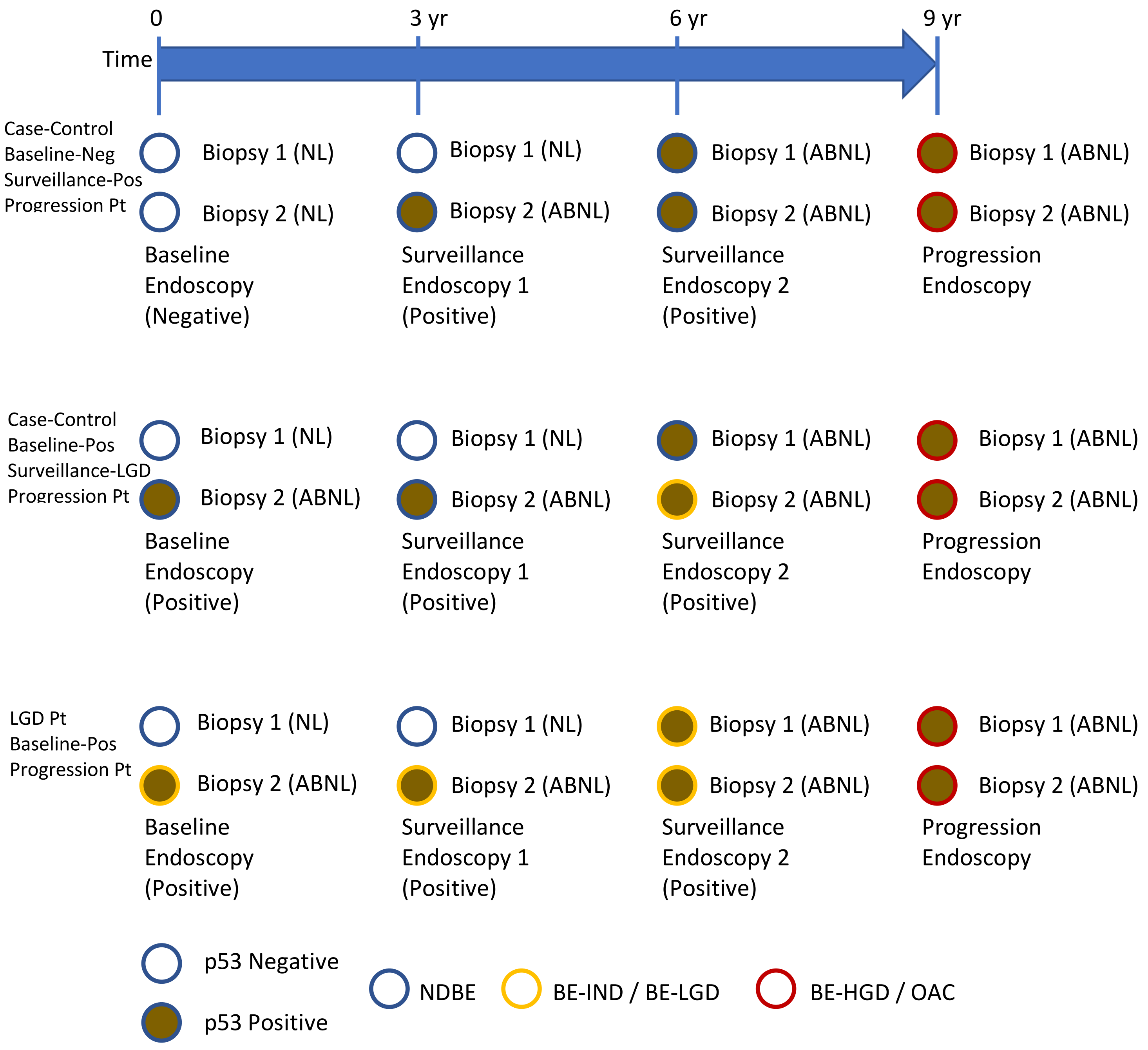

**Supplementary Figure S2. Accession timeline chart of baseline NDBE p53-NL progressors with follow-up NDBE biopsies prior to development of dysplasia.** Multiple cases had a subsequent p53-ABNL sample.

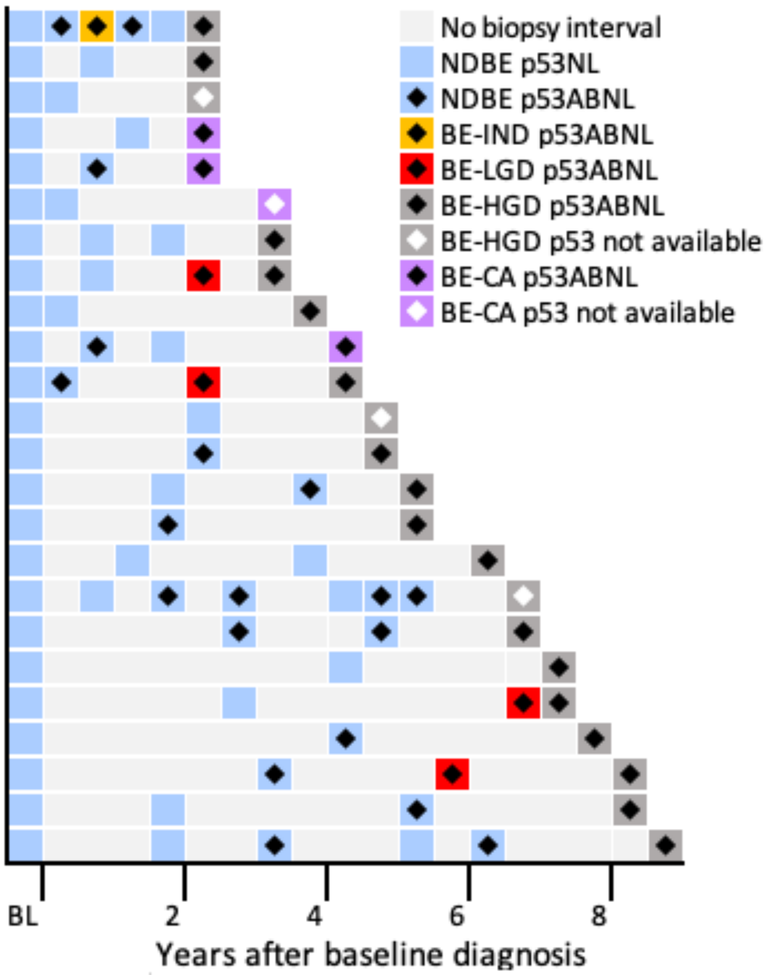

**Supplementary Figure S3. p53-ABNL vs time to progression.**

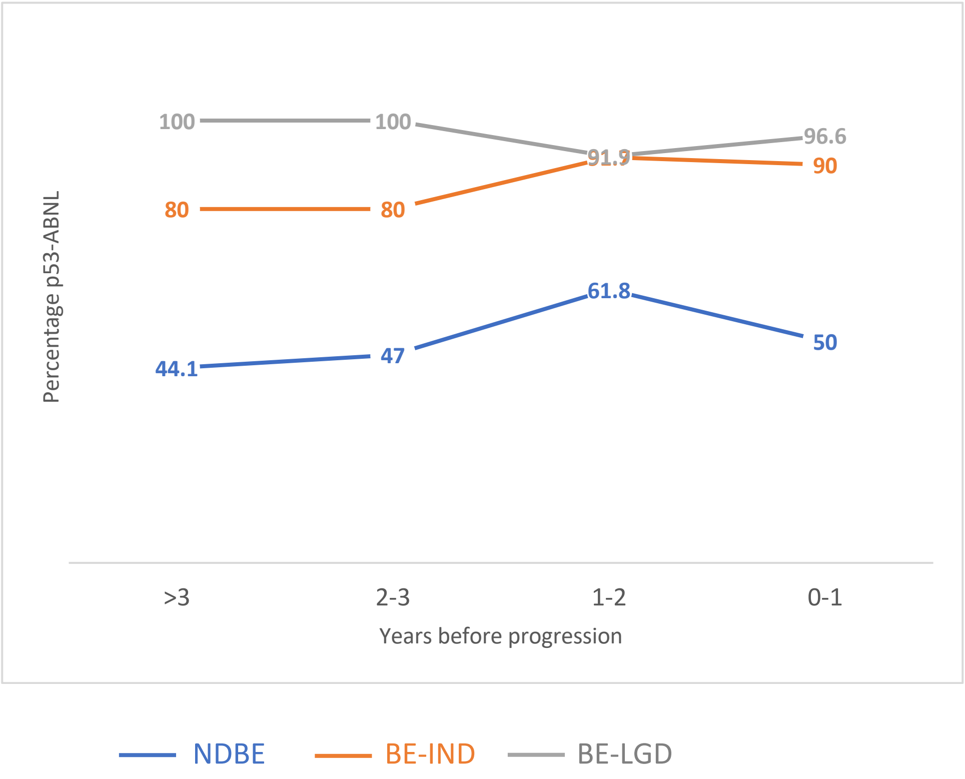

**Supplementary Figure S4. Timeline charts of subjects with intramucosal (A) and invasive carcinoma (B).** Multiple patients had progression from NDBE to invasive cancer within the recommended surveillance interval without a diagnosis of dysplasia.

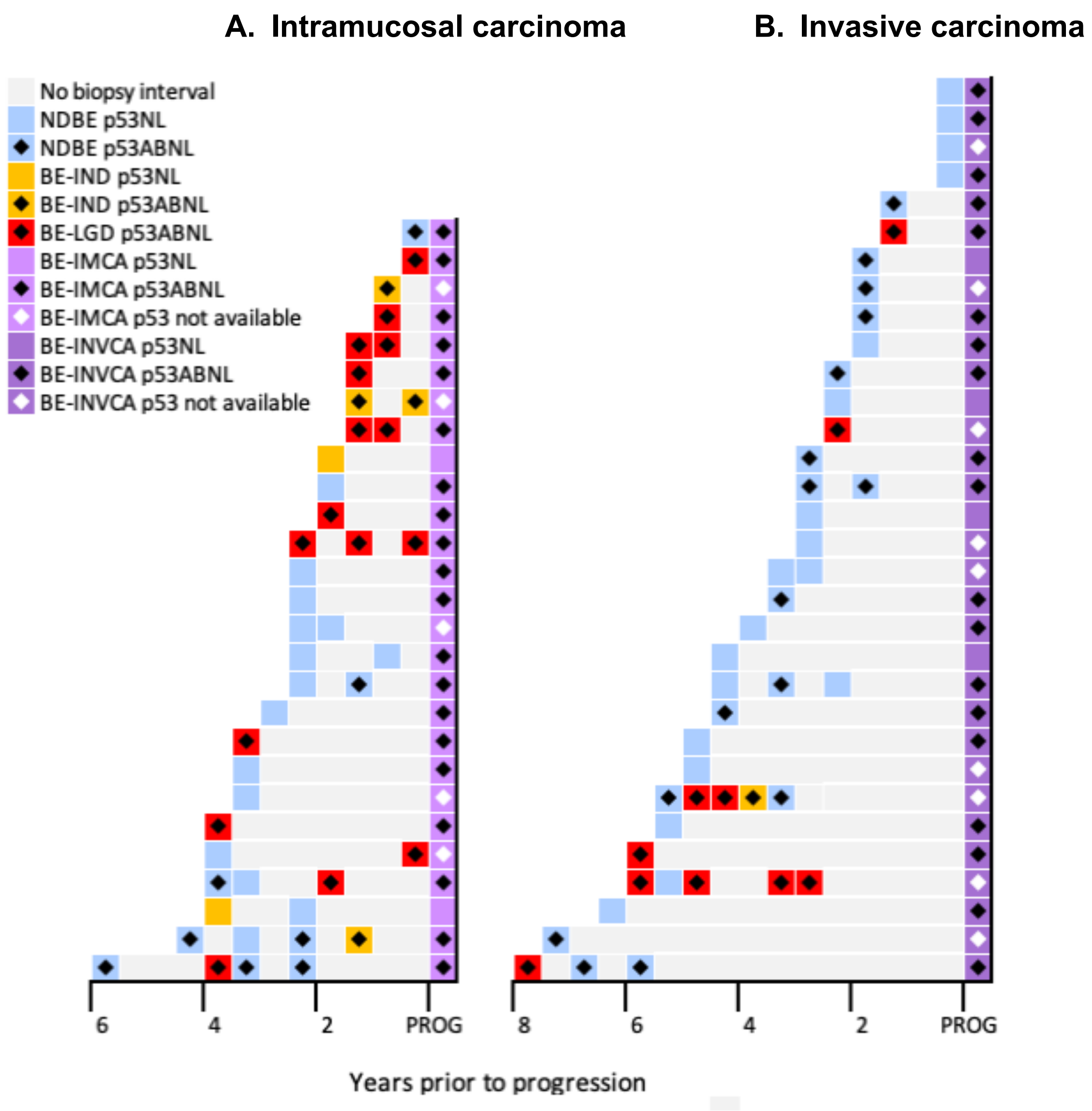

**Supplementary Figure S5. p53 IHC scoring** A) example of cells with negative p53 IHC (blue arrow), 1+ staining (tan arrow), 2+ staining (light brown arrow), and 3+ staining (dark brown arrow). B) example of wild type staining. C) Example of positive nuclear staining. D) Example of absent staining (positive). Glands quantified in Blue circles.

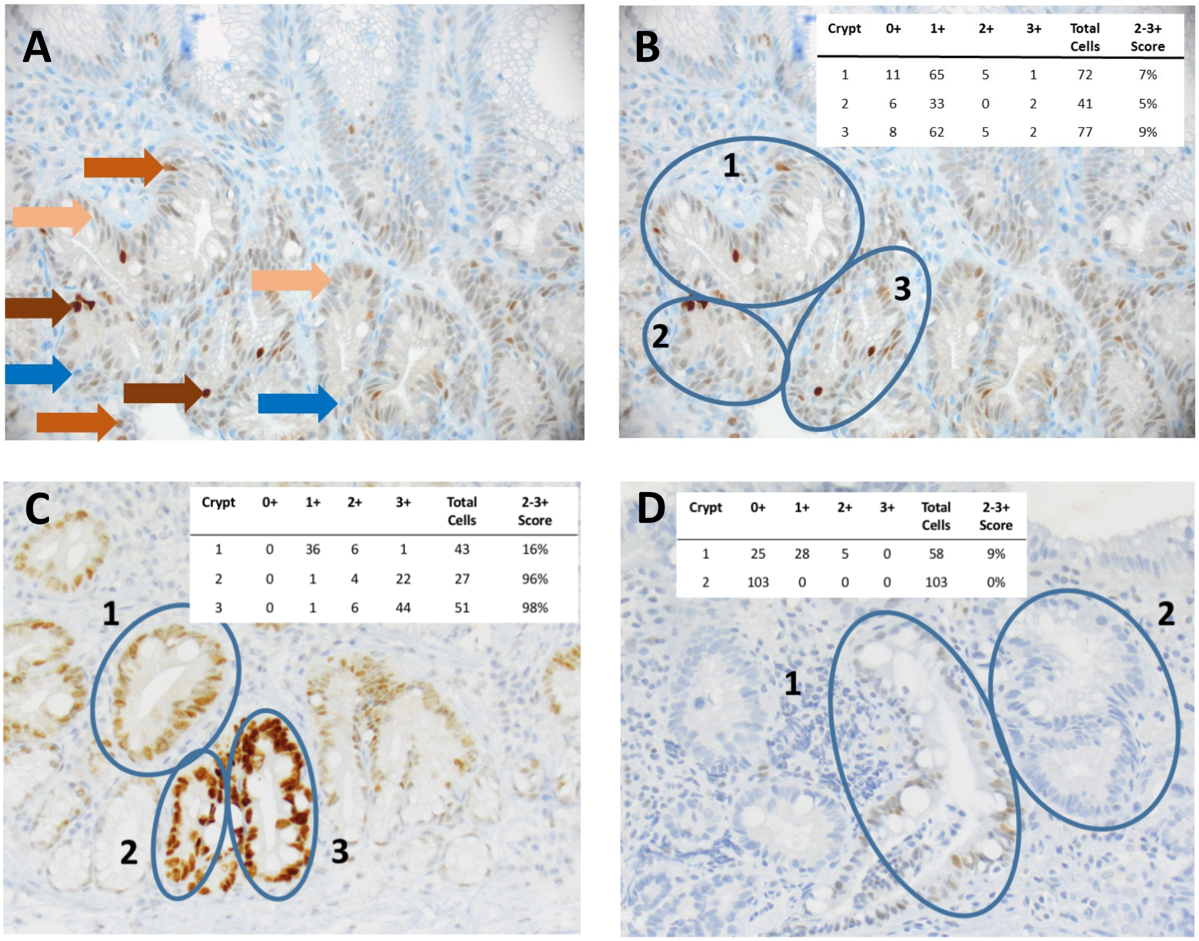

B

| Crypt | 0+ | 1+ | 2+ | 3+ | Total Cells | 2-3+ Score |
|---|---|---|---|---|---|---|
| 1 | 11 | 65 | 5 | 1 | 72 | 7% |
| 2 | 6 | 33 | 0 | 2 | 41 | 5% |
| 3 | 8 | 62 | 5 | 2 | 77 | 9% |

C

| Crypt | 0+ | 1+ | 2+ | 3+ | Total Cells | 2-3+ Score |
|---|---|---|---|---|---|---|
| 1 | 0 | 36 | 6 | 1 | 43 | 16% |
| 2 | 0 | 1 | 4 | 22 | 27 | 96% |
| 3 | 0 | 1 | 6 | 44 | 51 | 98% |

D

| Crypt | 0+ | 1+ | 2+ | 3+ | Total Cells | 2-3+ Score |
|---|---|---|---|---|---|---|
| 1 | 25 | 28 | 5 | 0 | 58 | 9% |
| 2 | 103 | 0 | 0 | 0 | 103 | 0% |

**Supplementary Figure S6. Distribution of p53 IHC positivity in NDBE biopsies**

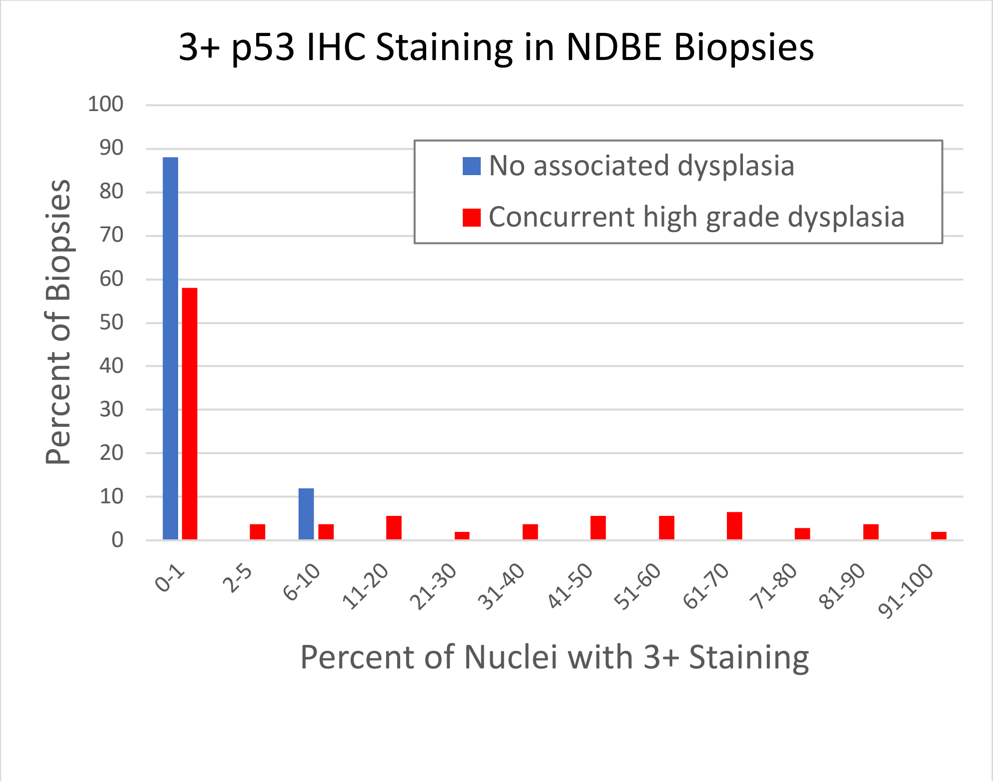

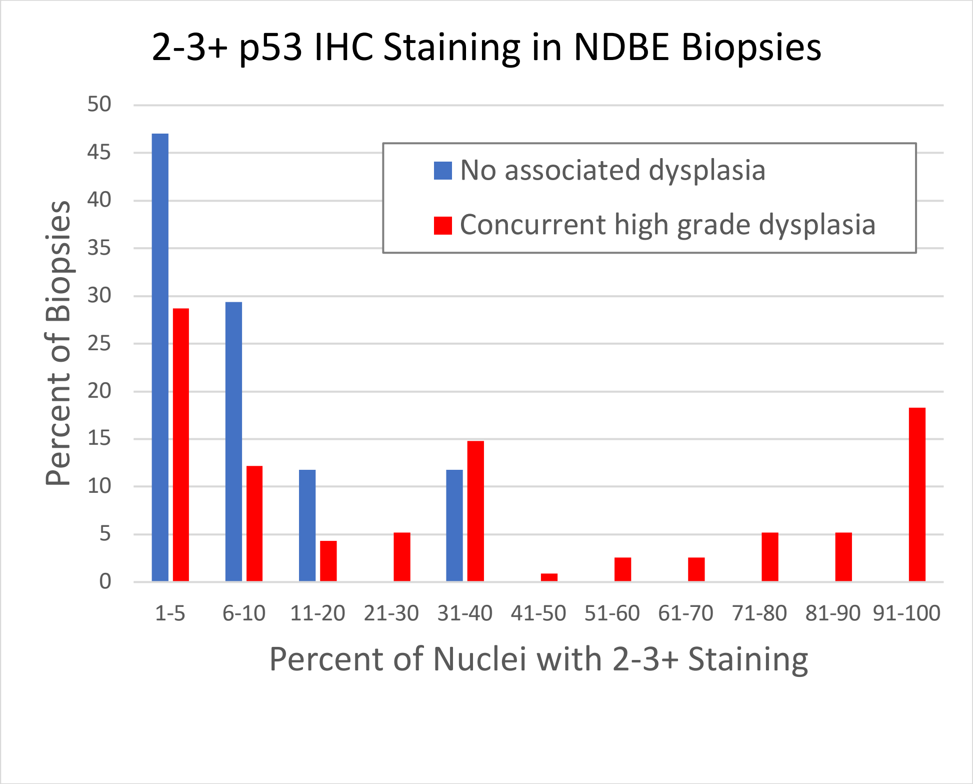

**Supplementary Figure S7. Kaplan-Meier analysis of progression free survival in patients with screening or surveillance baseline endoscopies.**

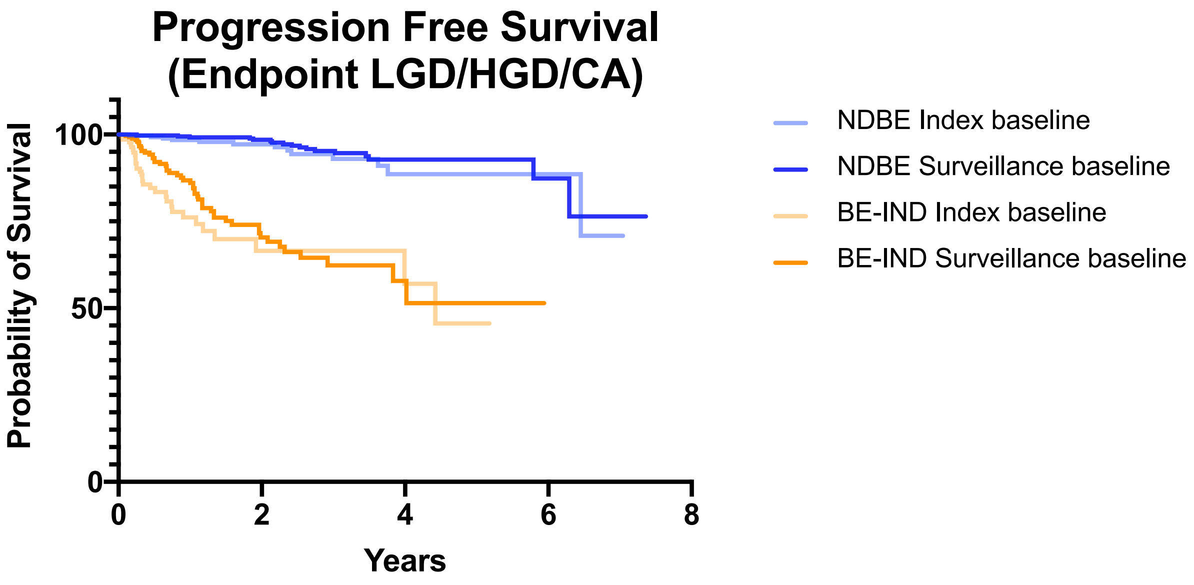

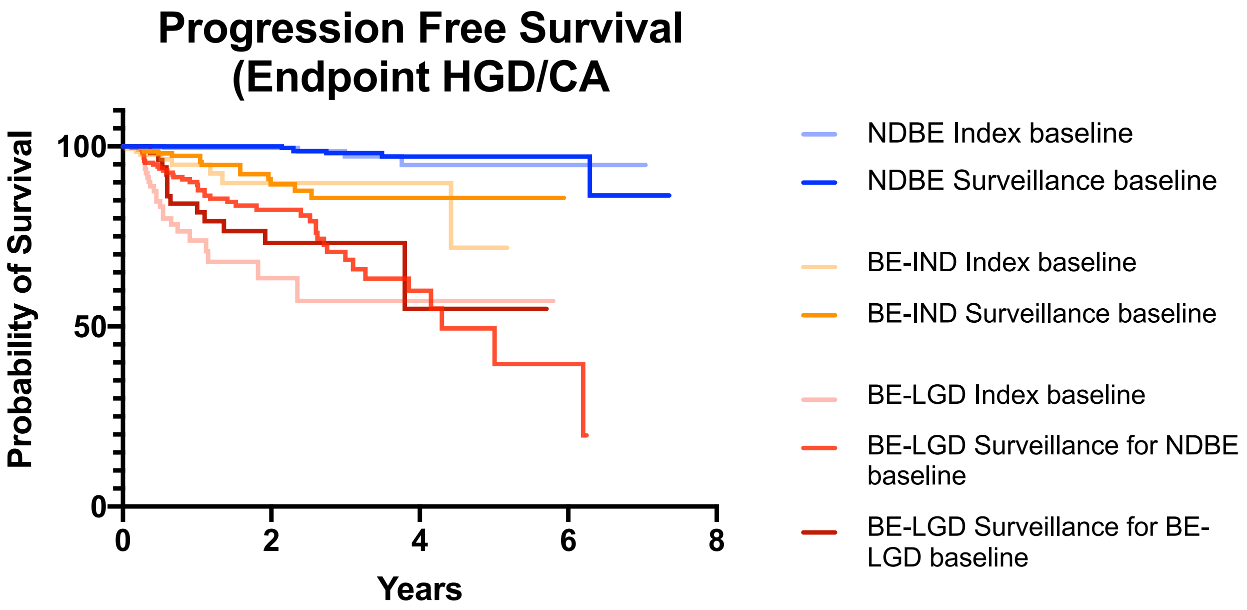
